# Epidemiology of sleep disorders during COVID-19 pandemic: A systematic scoping review

**DOI:** 10.1101/2020.10.08.20209148

**Authors:** Samia Tasnim, Mariya Rahman, Priyanka Pawar, Xinli Chi, Qian Yu, Liye Zou, Abida Sultana, E. Lisako J. McKyer, Ping Ma, Md Mahbub Hossain

## Abstract

**Background:** A growing burden of mental health problems has become a global concern amid the coronavirus disease (COVID-19) pandemic. Sleep disorders are major mental health problems associated with increased psychosocial stressors; however, no research synthesis is available on the epidemiology of it. In this systematic scoping review, we aimed to assess the current evidence on the epidemiological burden, associated factors, and interventions from the existing literature.

**Method:** Seven major health databases and additional sources were searched to identify, evaluate, and synthesize empirical studies on the prevalence and correlates of sleep disorders and available interventions. The Joanna Briggs Institute Methodology for Scoping Review were used, and the findings were reported using the Preferred Reporting Items for Systematic Reviews and Meta-Analyses extension for Scoping Reviews (PRISMA-ScR) checklist.

**Results:** A total of 78 articles were retrieved, the prevalence of sleeping disorders ranged from 2.3% to 76.6%. Age, sex, level of education, physical and mental health, COVID-19 related factors, occupation especially being health care workers (HCW) were the main associated factors. Only two intentions were identified to address the issue.

**Conclusion:** The finding of this review indicated a high burden of sleep disorder with limited interventions that necessitate informing policymakers and practitioners to facilitate future research and implementations.

**Brief summary:** *Current Knowledge/Study Rationale:* Despite the paramount importance of sleep for the physical and mental wellbeing of individuals, sleep hygiene is often neglected which resulted in a high prevalence of sleep disorders across the globe. This condition is likely to worsen amid this pandemic. This is the first systematic scoping review of sleep disorders during the COVID-19 pandemic.

*Study Impact:* The findings of our study suggest a high prevalence of sleep disorder and highlight a wide range of socio-demographic factors to identify population groups vulnerable to the adverse outcomes of sleep disorder with limited interventions. These pieces of evidence will guide clinicians to make informed choices for better management of patients and aid public health professionals to prevent sleep disorder epidemic concurring with the current pandemic.

## 1. Introduction

Novel coronavirus disease or COVID-19, an acute respiratory illness caused by a newly discovered SARS-CoV-2 virus emerged in December 2019 ^1,2^. Since then, it has rapidly surged to Europe, especially Italy and Spain, the USA, and progressed to become a global pandemic. In the last couple of months, it’s reported to be spreading to the countries in other parts of Asia, Africa, and Latin America. Globally, by September 27th, 2020, a total of 32,730,945 cases of COVID-19 has been reported, including 991,224 deaths ^3^. Of note, the World Health Organization declared it a global pandemic on March 11th, and after the declaration, most of the countries worldwide entered nationwide lockdown to prevent the spread of the infection ^4^. While the clinical care practitioners and public health experts were focused on containing the spread of the virus, the COVID-19 pandemic and related quarantine measures have taken a heavy toll on people’s mental health^4,5^. Historically quarantine has been related to anxiety, depression, panic, irritability, somatic disorder, and insomnia^6–8^. Moreover, a high level of stress and trauma-related disorders are byproducts of being isolated^5,9,10^. Moreover, factors like an extended period of isolation, fear of infection, uncertainty, disappointment, fatigue, stigma, inadequate data and information regarding the disease, insufficient supplies, and economic damage also negatively impact individuals’ psychological wellbeing^11^. Psychosocial stressors like anxiety, stress, altered lifestyle with little to no social support, and fear may affect the pattern of sleep among individual often leading to sleep disorders ^12,13^.

Sleep is an essential physiological activity in keeping up with physical and mental wellbeing and better life quality^14,15^. Breach in the normal sleep cycle can lead to insufficient sleep and prolonged alertness, hence increasing the event of insomnia, nightmares, daytime instability, and fatigue^14^. Recent studies have shown that sleep disorders impact up to 1 in 4 adults^16^. They are also found to be associated with a wide range of adverse health outcomes, for example, increased risk of obesity, diabetes, hypertension, cerebrovascular diseases, malignancy, musculoskeletal diseases, septicemia, and metabolic syndrome^14,15,17^. Further evaluations have found that 50-70 million adults in the United States have at least one sleep disorder^16^, while the prevalence of sleep disorders among the Australians and Netherlanders are 20-35% ^18^and 27.3% ^17^respectively. Potential risk factors of sleep disorders include severe stressful circumstances, depression, anxiety, trauma, low socioeconomic condition, urban living, increased use of technology, and social media^14,16,17^.

A substantial amount of evidence suggests a high prevalence of various forms of sleep disorders in the global community. This situation is likely to worsen in the current circumstances with numerous psychological stressors. Many studies have reported a growing burden of a wide range of mental disorders, including sleep disorders^19^. However, to the best of our knowledge, there is a scarcity of concrete evidence on the magnitude of sleep disorders among individuals affected by this pandemic. As there is a growing concern on mental health problems during COVID-19 research synthesis may play a critical role in understanding the burden of those problems addressing the same^6,19^. This scoping review aims to address this knowledge gap through systematically evaluating the current evidence on the epidemiological burden of sleep disorders, associated factors, and interventions addressing the problems. The specific questions for this scoping review are listed below:

- What is the epidemiological burden of sleep disorders among different populations during COVID-19?
- What are the factors associated with sleep disorders during COVID-19?
- What are the available interventions for addressing sleep disorders amid COVID-19?

## 2. Methods and materials

This scoping review was conducted using the Joanna Briggs Institute (JBI) methodology for scoping reviews^20^. Moreover, the findings of this review are reported using the Preferred Reporting Items for Systematic Reviews and Meta-Analyses extension for Scoping Reviews (PRISMA-ScR) checklist^21^. The protocol of this scoping review has been registered with The Open Science Framework^22^.

### 2.1 Search strategy

We searched MEDLINE, Embase, Academic Search Ultimate, Cumulative Index to Nursing and Allied Health Literature (CINAHL), Web of Science, and APA PsycInfo databases using the keywords with Boolean operators “AND/OR” as mentioned in Table 1. The search was first conducted on May 13^th^, 2020 and updated on August 12^th^, 2020.

**Table 1:** Search terms for this scoping review.

| **Search terms** | **Keywords (searched within titles, abstracts, and subject headings fields)** |
| --- | --- |
| **1** | “COVID-19” OR “2019-nCoV” OR “2019 coronavirus” OR “Wuhan coronavirus” OR “Wuhan virus” OR “Wuhan pneumonia” OR “2019 novel coronavirus” OR “novel coronavirus” OR “SARS-CoV-2” |
| **2** | “sleep” OR “sleep disturbances” OR “sleep disorders” OR “sleep problems” OR “insomnia” OR “circadian rhythm” OR “restless leg syndrome” OR “sleep apnea” OR “narcolepsy” OR “daytime dysfunction” OR “daytime sleepiness” |
| **3** | prevalence OR incidence OR epidemiology OR frequency OR case OR rate OR occurrence OR correlates OR determinants OR predictors OR "risk factors" OR "associate factors" OR interventions OR treatment OR therapy OR management |
| **Final search strategy** | 1 AND 2 AND 3 |

The keywords were searched within the titles, abstracts, and subject headings fields. Moreover, as COVID-19 literature started to get published since late 2019, we limited our search within 2019 and 2020. Moreover, we searched the reference lists and citing articles in Google Scholar to identify additional articles that could have met our criteria.

### 2.2 Inclusion criteria

#### 2.2.1 Participants

In this scoping review, we included participants irrespective of their sociodemographic conditions. This makes our review inclusive for all types of participants who fulfill remaining criteria of this review.

#### 2.2.2 Concepts

This review focused on sleep disorders, which can be defined by the International Classification of Diseases or Diagnostic and Statistical Manual of Mental Disorders ^23,24^. Moreover, sleep abnormalities expressed as insomnia, excessive sleepiness, poor sleep quality, and abnormal events that occur during sleep will also be considered as sleep disorders in this review ^25^. Studies reporting the prevalence, incidence, frequency, score, level or any forms of quantitative assessment of sleep-related conditions were included in this review.

#### 2.2.3 Context

This review especially emphasized on COVID-19 as the context. Therefore, studies conducted among populations affected by COVID-19 (doctors or patients) or population at risk (general population who could have had infected with COVID-19) were be considered in this review. Moreover, studies without mentioning relevance to COVID-19 were excluded from this review.

#### 2.2.3 Types of sources

This review included original studies, cross-sectional or longitudinal in nature, published as peer-reviewed journal articles. Studies published in English language were included in this review. Therefore, unpublished works, non-original articles (for example, letters with no original research reports, editorials, reviews, commentaries etc.), non-peer reviewed articles, and studies in languages other than English were excluded from this review.

#### 2.2.4 Study selection

After searching the databases, all the citations were imported to Rayyan QCRI, a cloud-based software for systematic reviews^26^. Two authors (ST and MR) independently evaluated those citations using the inclusion criteria of this review as stated earlier. At the end of independent screening, potential conflicts were reviewed and resolved based on discussion with a third author (MMH). Finally, the full texts of the included citations were assessed and articles meeting all the criteria were considered for data extraction. A flow chart of the study selection process is depicted in Figure 1.

**Figure 1:**
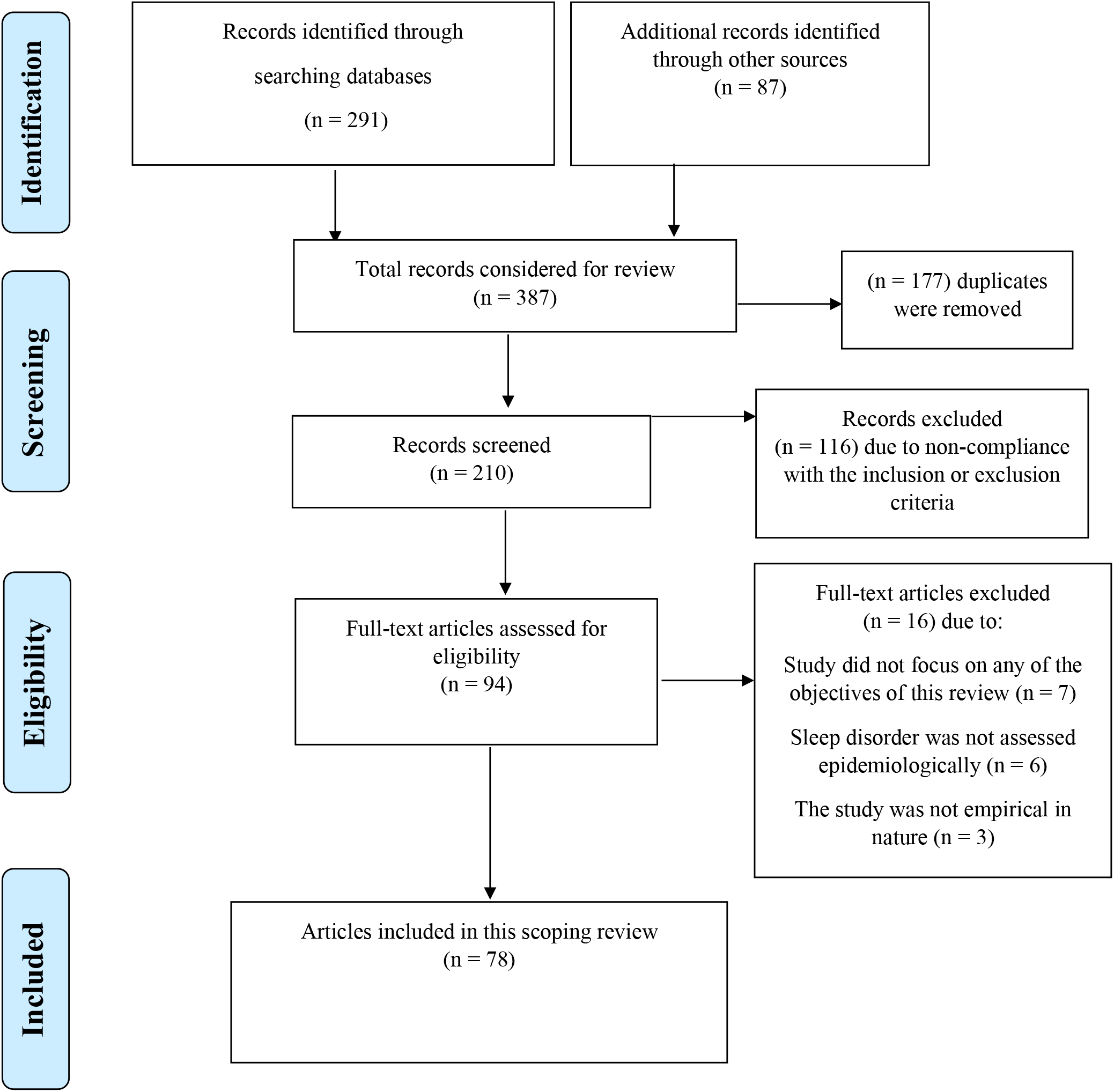
Flow diagram of the systematic scoping review.

### 2.3 Data extraction

A data extraction form was prepared as shown in Table 2. Two authors (ST and MR) independently extracted data using this checklist. At the end of this phase, data for each article were reviewed by a third author to check consistency and potential conflicts were re-assessed by three reviewers and a consensus was made based on discussions. The finalized dataset was reviewed independently by two more co-authors (PP and AS) before final synthesis.

**Table 2:**
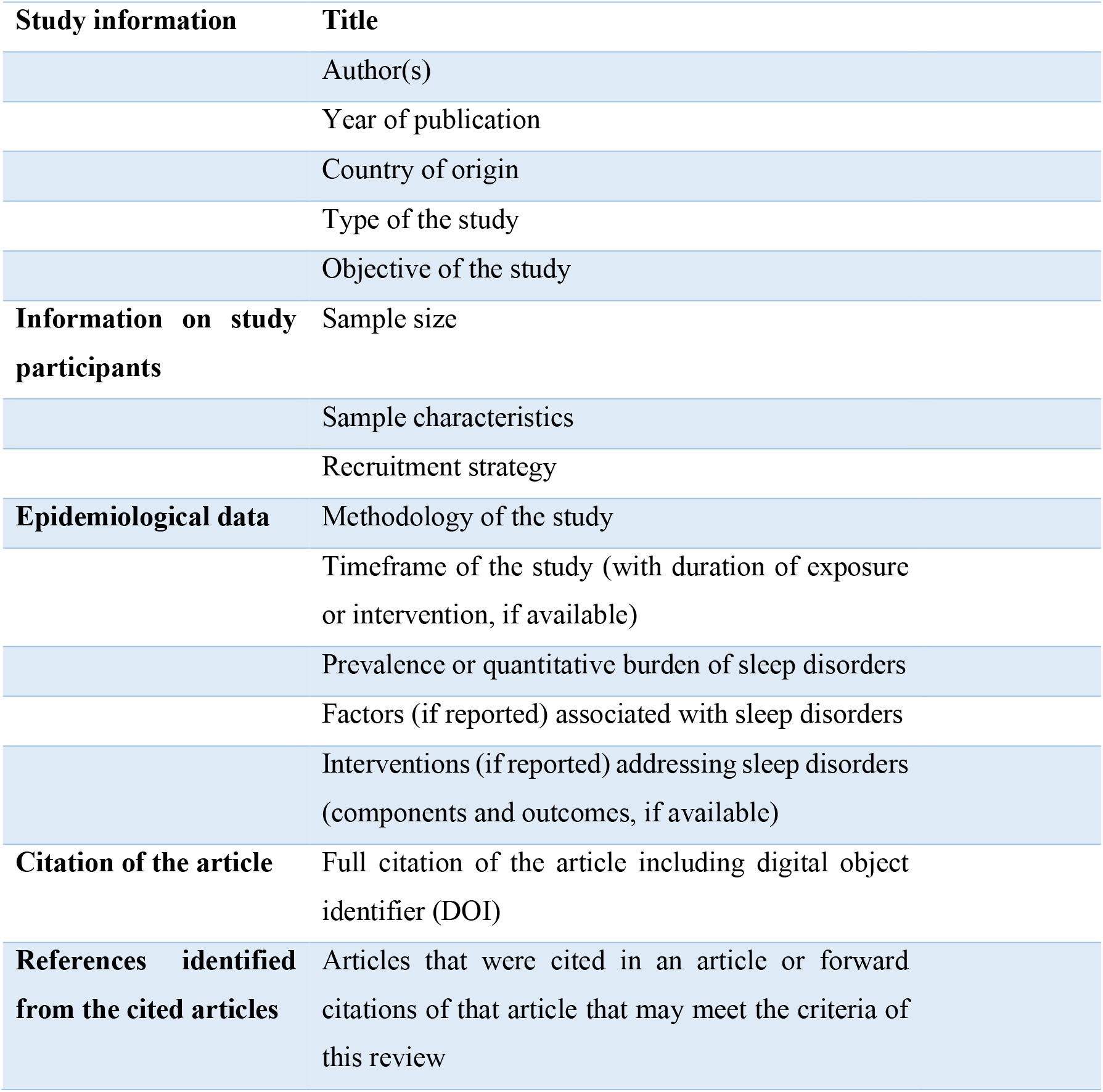
Data extraction instrument.

### 2.4 Data presentation

Data extracted from the included articles were narratively synthesized and presented using tables and a commentary on key findings on the study characteristics, samples, and epidemiological findings as the quantitative burden and associated factors of sleep disorders during COVID-19 and interventions addressing the same. As per the JBI methodology, scoping reviews do not aim to evaluate the quality of the studies. Therefore, no quality evaluation was done in this review.

## 3. Result

We found 291 citations from searching the databases and 87 citations from additional sources, totaling 387 citations. After removing 177 duplicates, titles and abstracts of 210 articles were evaluated and 94 articles met the pre-set inclusion criteria. From the remaining articles 16 articles were removed after evaluating the full text. Finally, 78 articles met all criteria and were included in this scoping review.

### 3.1 Characteristics of the included studies

#### 3.1.1 Study design

Most of the studies 82% (n = 64) were of cross-sectional design. For example, Zhang et al., 2020 conducted cross-sectional survey among the hospital workers in China. A few included studies (n = 6) were of cohort studies. For example, the study by Nalleballe et al., 2020^27^ reported the findings from their cohort of patients of COVID-19. Remaining were case-control (n = 4) and pre-post study (n = 3) design.

#### 3.1.2 Setting

Almost half of the studies (n = 36, 46%) were based on community setting. For example, the study by Gao et al.,^28^ recruited 699 American adult participants to find out the effects of the pandemic related lockdown on their mental health. However, other half of the studies (n = 37, 47%) recruited participants from hospitals or clinics. The study by Amerio et al.,^29^ included general practitioners working in Genoa, Italy evaluate the mental health effects of the Covid-19 pandemic on the healthcare workers.

#### 3.1.3 Geographical scope

One third of the studies, (n = 25, 32%) were conducted in urban areas. For example, 76.5% of the participants recruited in the study by Voitsidis et al.,^30^ were urban citizens. Similarly, the study by Xiao, Zhang, Kong, Li, & Yang et al.,^31^ reported 89.4% participants were recruited from urban area. In 13.75% (n = 11) of the studies participants lived equally in urban and rural areas. For example, Renzo et al., ^32^ reported that the percentage of participants living in rural and urban areas were comparable. However, in a few studies (n = 18, 23%) participants also came from small towns or rural areas.

#### 3.1.4 Country

47.5% (n = 38) of the studies were conducted in China. The study by Fu et al.,^33^ was based on Chinese population where they recruited participants from Wuhan, China. 8.75% (n = 7) of the studies were based on USA. Wright et al.,^34^ was an observational study based on the university students of the USA. 8.75% (n = 7) of the studies were based on Italy. For example, the study by Gualano et al.,^35^ was based on Italy.

#### 3.1.5 Sample

A large variation was noted in the sample size of the included studies. The sample size ranged from 26 to 40,469. Studies that provided intervention or conducted in-person assessments had a significantly smaller sample size. For example, Liu et al., ^36^ conducted a randomized control trial of progressive muscle relaxation for insomnia among the COVID-19 patients in a hospital setting and had a small sample size of 51 participants. Similarly, Korkmaz et al.,^37^ conducted face to face evaluation of several mental health conditions of health care workers employed in service for COVID-19 and had a sample size of 140 participant only.

However, studies that utilized the online surveys to collect data had a relatively larger sample size. For example, S. J. Zhou et al.,^38^ collected data from 11,835 participants in China via online survey forms.

#### 3.1.6 Sampling technique

65% of the studies (n = 51) mentioned specific sampling techniques. Among them, 35% of the studies (n = 27) used the convenient sampling technique to recruit the participants. For example, Diomidous, et al.,^39^ used convenient sampling technique to include health care providers for their study. Some studies (n = 6) also used the random sampling technique. For example, Liu et al.,^36^ randomly selected 51 patients from the list of patients with confirmed COVID-19 admitted to the Hainan General Hospital for participating in their study. A few studies used clustered sampling technique. For example, Abdulah and colleagues’ ^40^ used the clustered sampling technique. They obtained a list of local physicians who work in different medical settings. The participants from one pediatric, one emergency, one special corona, and one maternity and gynecology hospital were invited to participate in their online survey.

#### 3.1.7 Disorders investigated

All the studies assessed insomnia/sleep disorder or quality of sleep. Most of the studies (n = 38, 49%) also investigated the level of anxiety among participants. For example, Mazza et al.,^41^ assessed prevalence of anxiety among the COVID-19 survivors in Italy. Depression/depressive symptoms were assessed by 88 studies. For example, Huang et al.,^42^ measured the burden of depression among Chinese public during the outbreak. Loneliness, suicidal ideation, somatic disorders, and Obsessive-Compulsive Disorder (OCD) were among other less frequently assessed disorders.

#### 3.1.8 Mode of data collection

85% of the studies (n = 66) collected data through online surveys. For example, W.D.S. et al.,^43^ collected data using online surveys on different social media platforms. Similarly, J et al.,^44^ collected data from the Chinese population using the WeChat (the most widely used mobile app). Only 7% of the studies utilized more than one means of data collection. For example, Renzo et al.,^32^ collected data using face to face interview, online survey and from reviewing hospital records of the patients. Remaining studies (n = 6, 7%) collected data solely from face to face interviews. For example, Türkoğlu et al.,^45^ collected data from in person evaluation of the children who participated in the study.

#### 3.1.9 Screening instruments and cut off value

Majority of the studies (n = 50, 64%) employed the PSQI for diagnosing the severity of the insomnia. For example, Huang & Zhao et al.,^42^ used the Chinese version of PSQI whereas, the study by Innocenti et al.,^46^ used the Italian version of PSQI among their participant. A little over half of the studies (n = 42, 53%) used the ISI scale for diagnosing insomnia. Gualano et al.,^35^ used ISI scale to diagnose insomnia among the Italian population. Few studies (n = 15,19%) also utilized the AIS scale for diagnosis. For example, Tselebis et al.,^47^ used AIS for diagnosing insomnia among COVID-19 survivors.

Majority of the studies used score of ≤ 7 for PSQI, ≤ 10 for ISI, and ≤ 6 for AIS scale as cut of value for diagnosing the severity of insomnia.

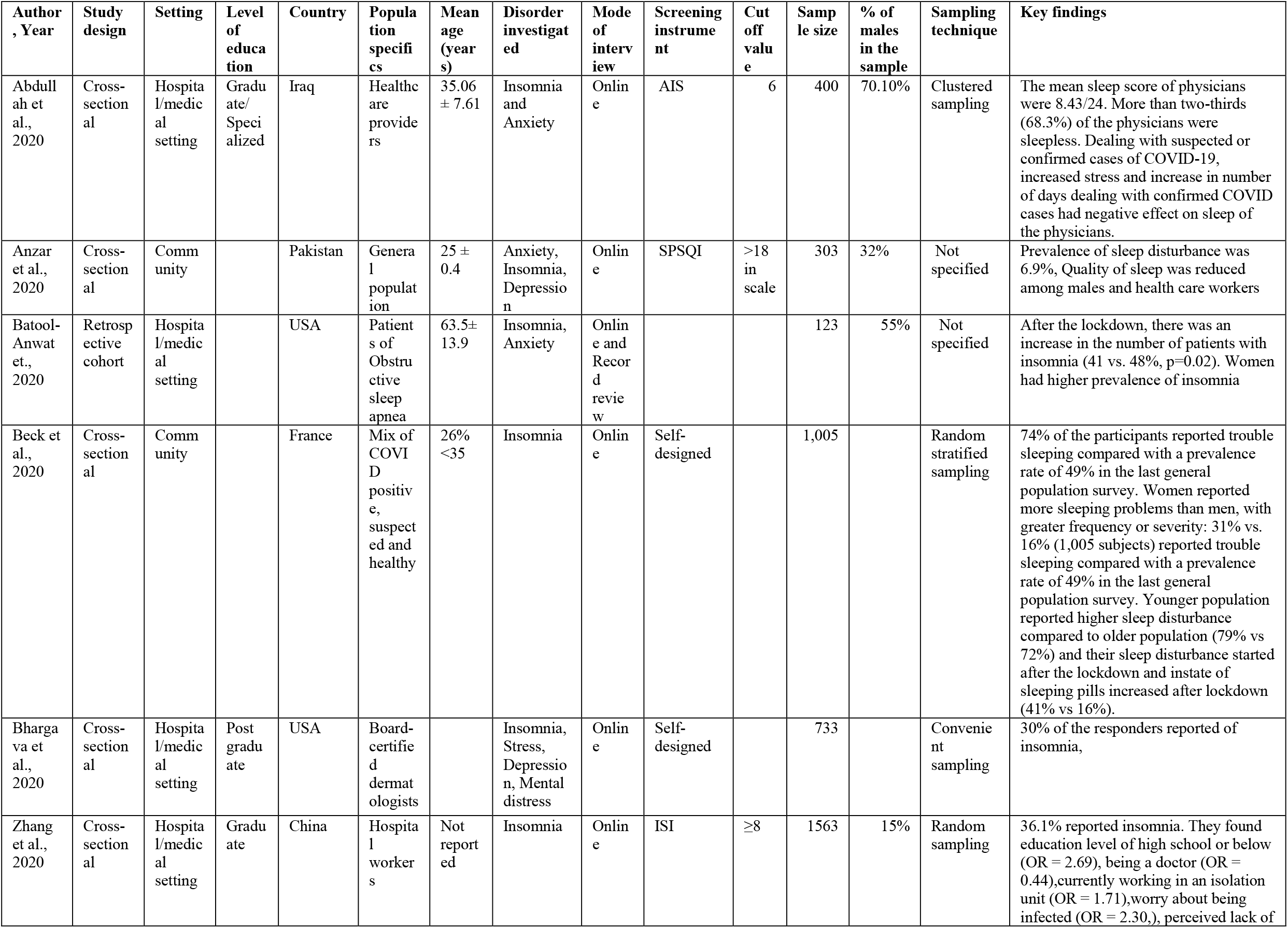
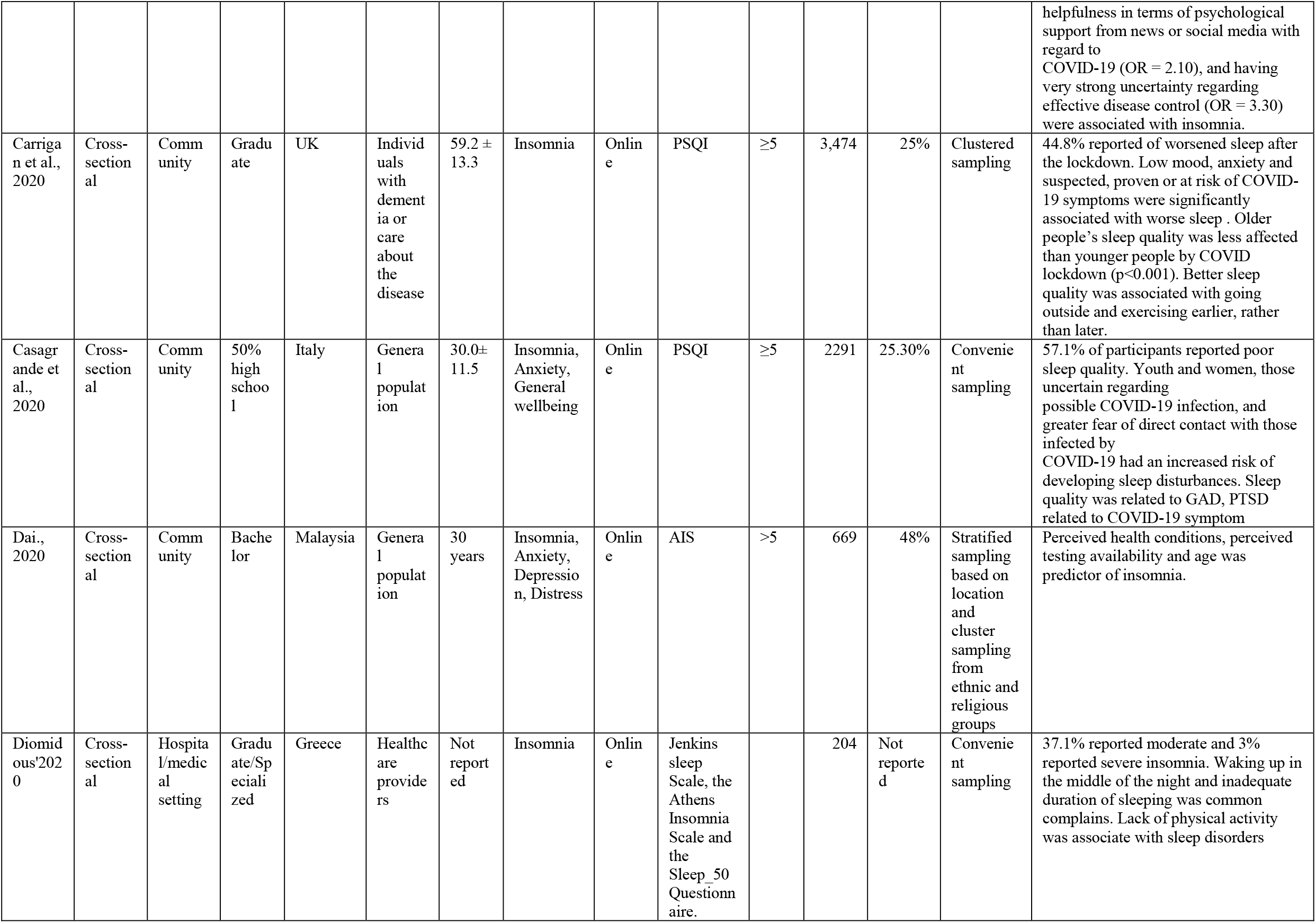
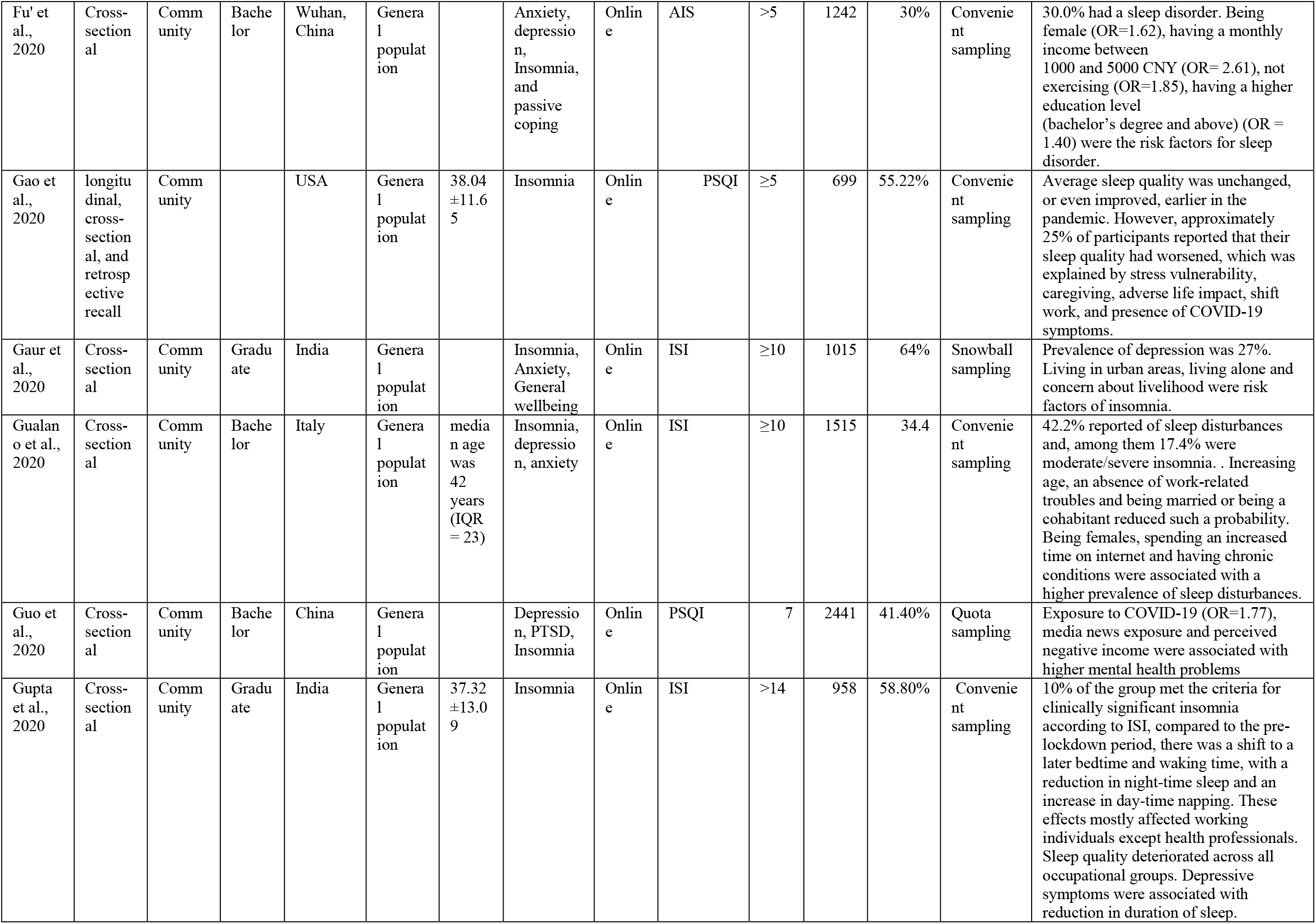
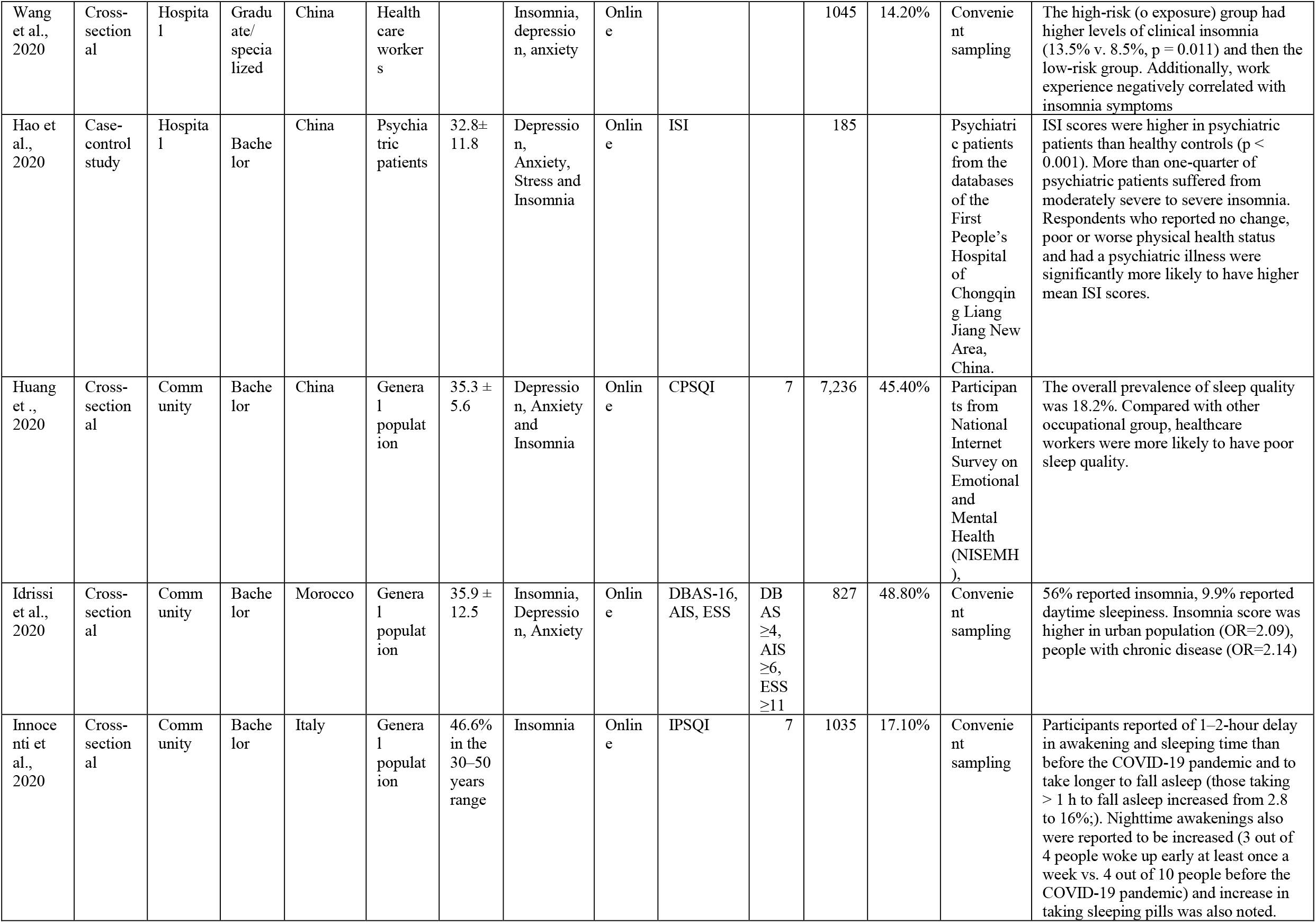
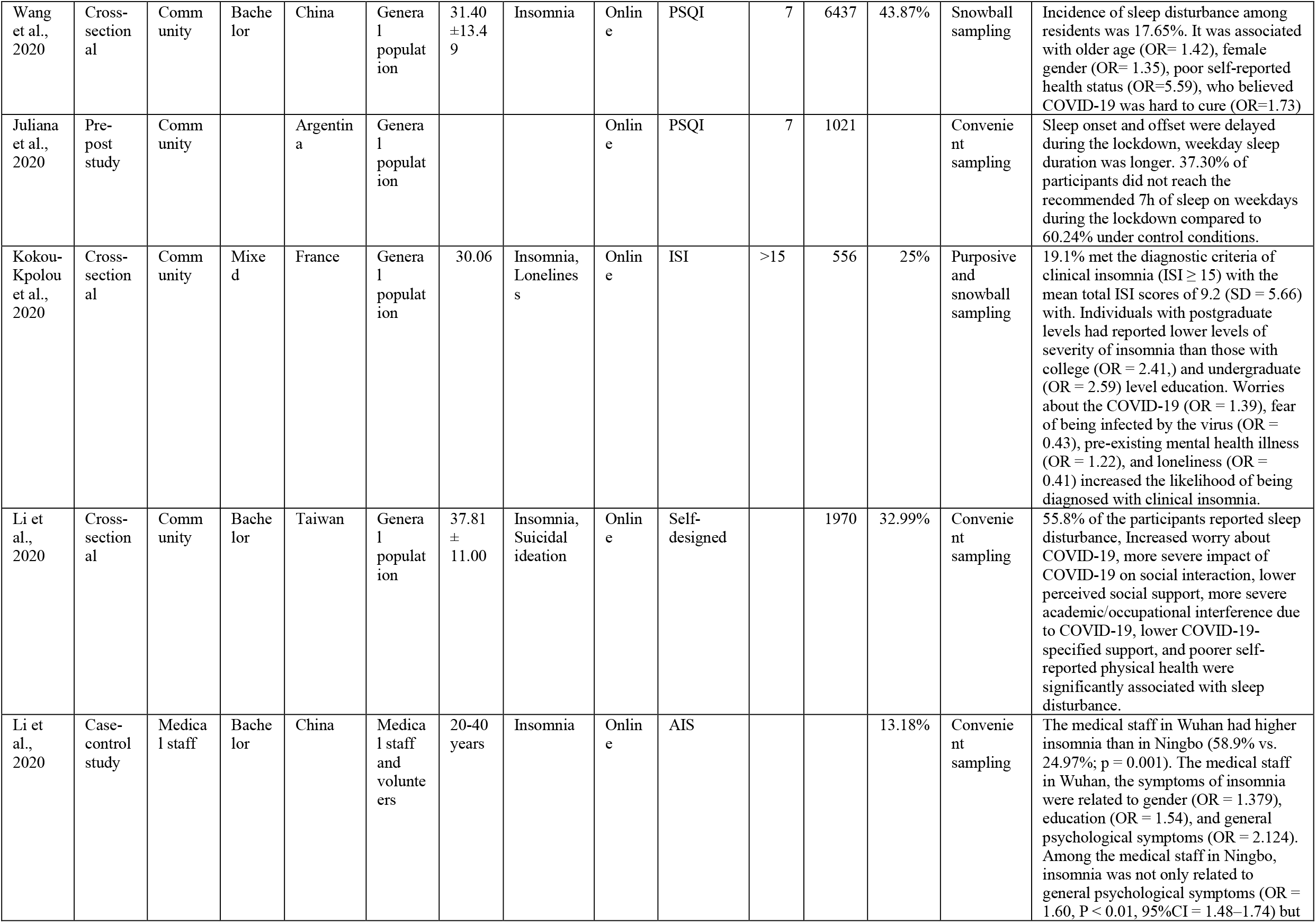
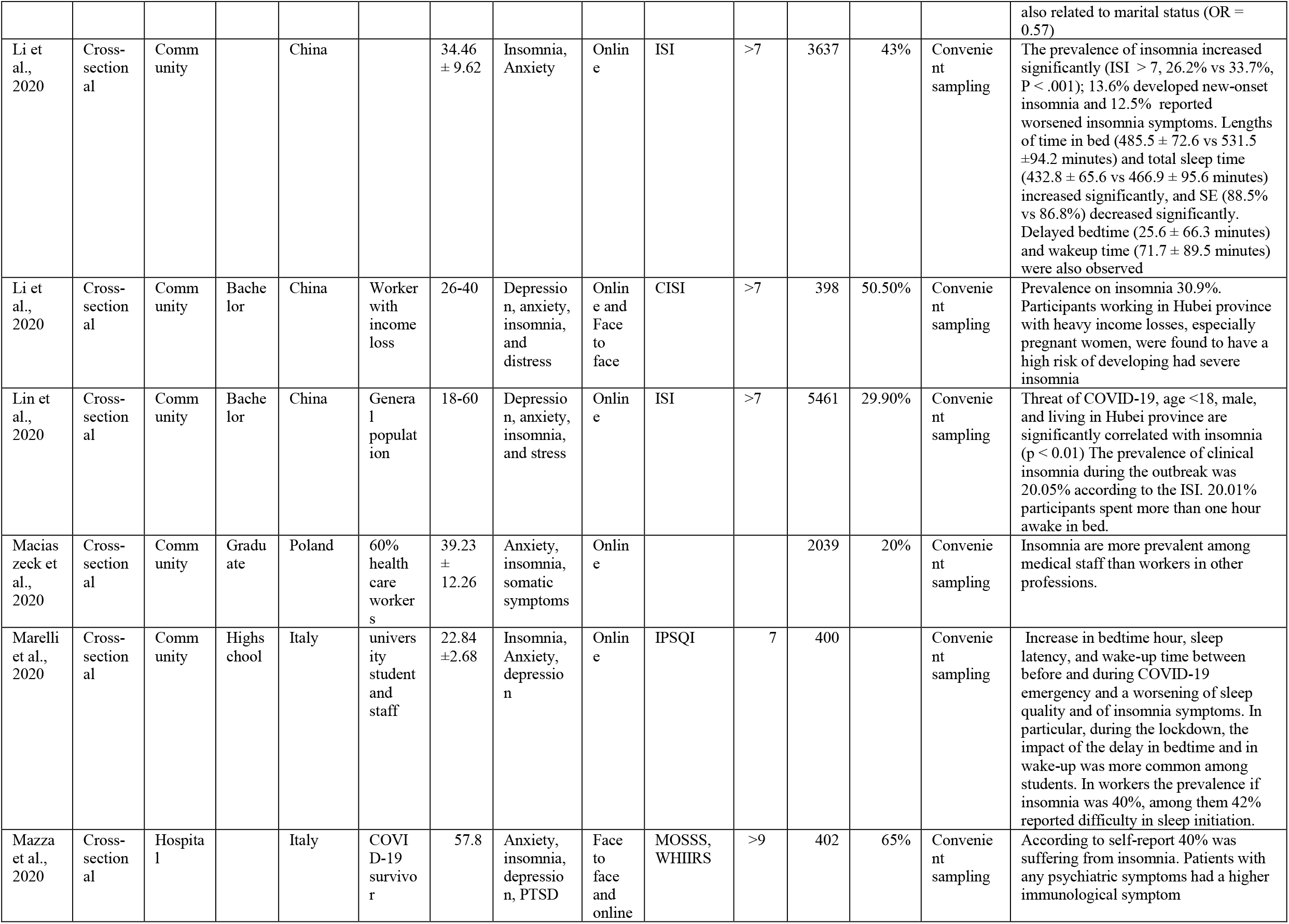
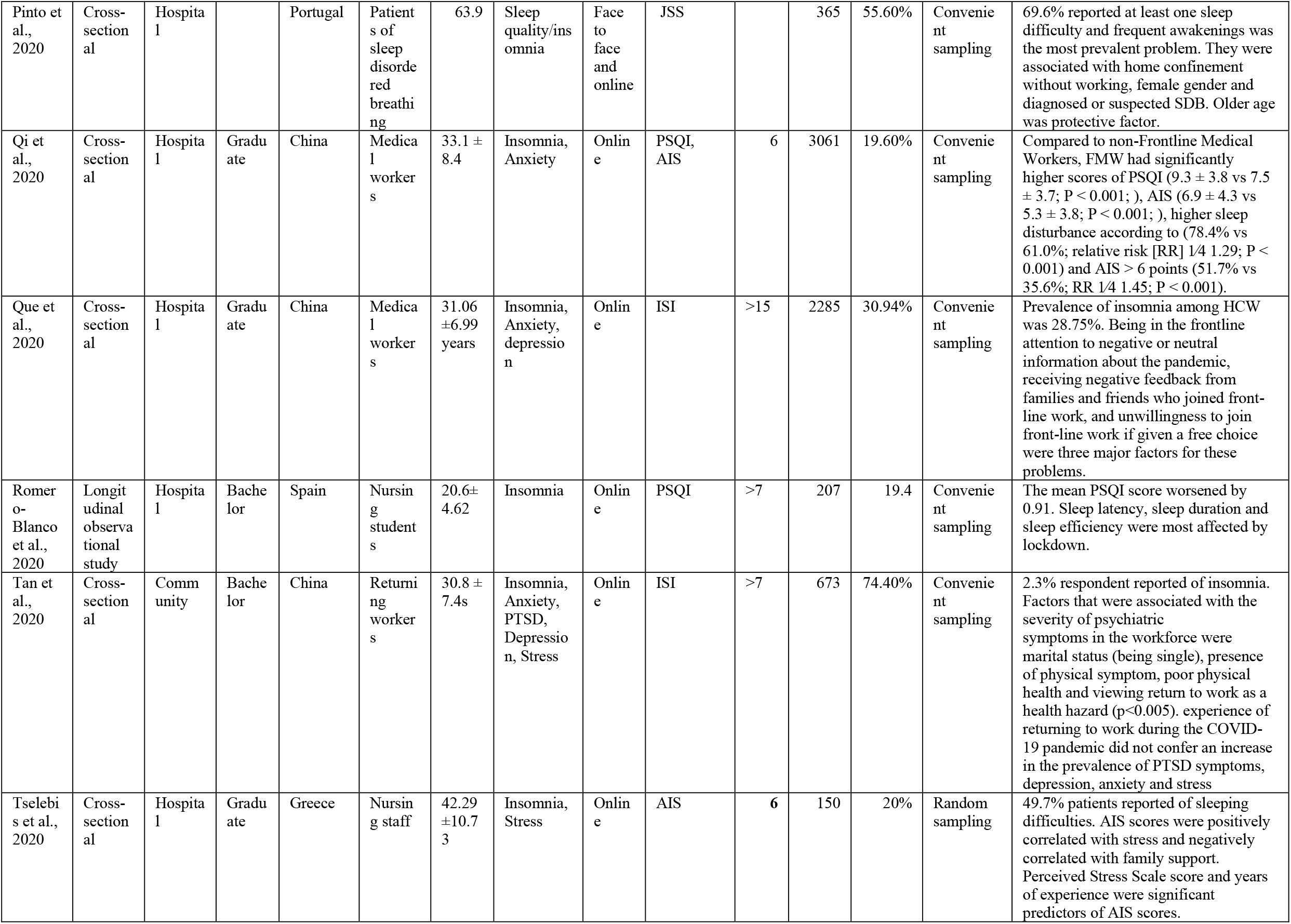
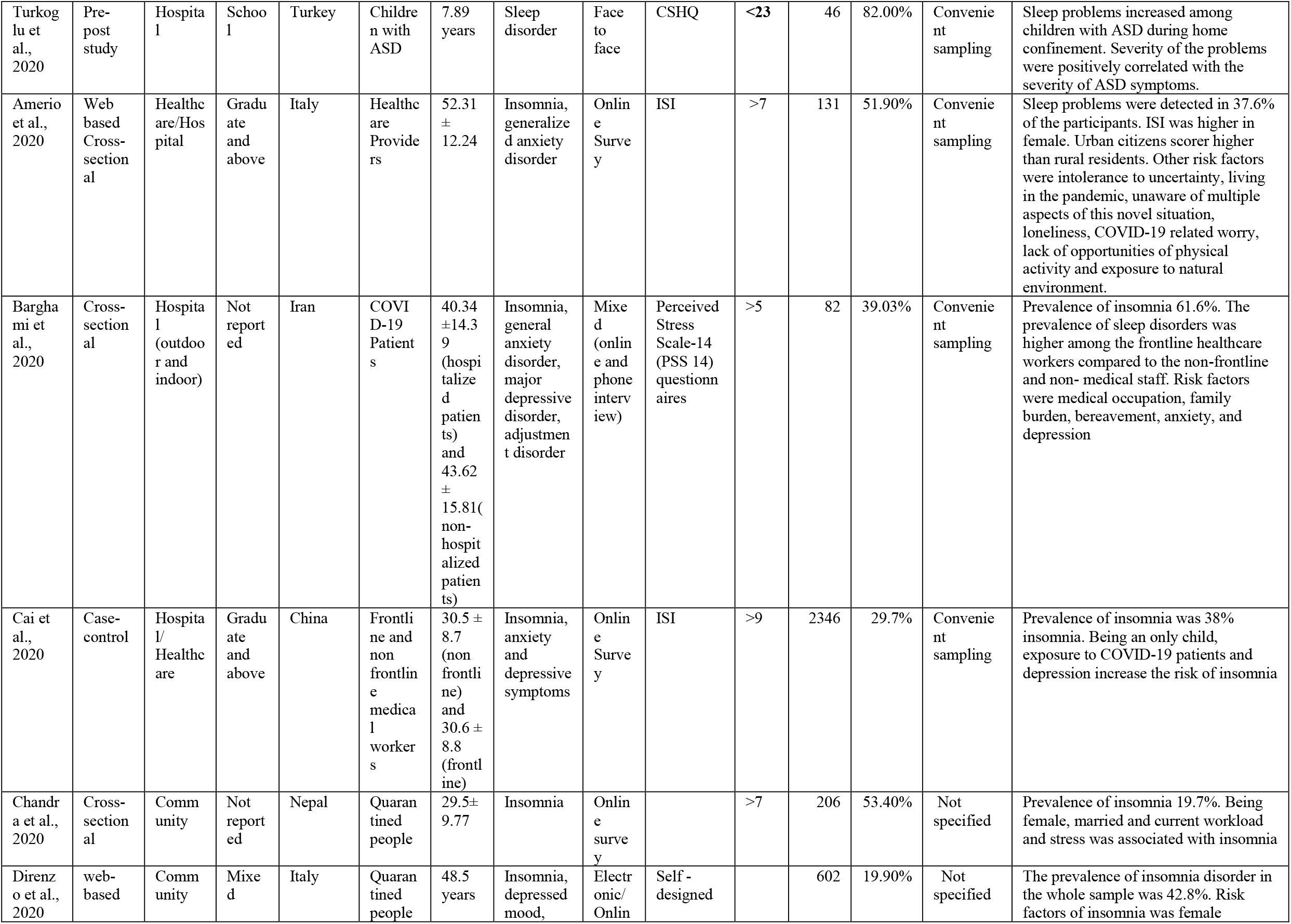
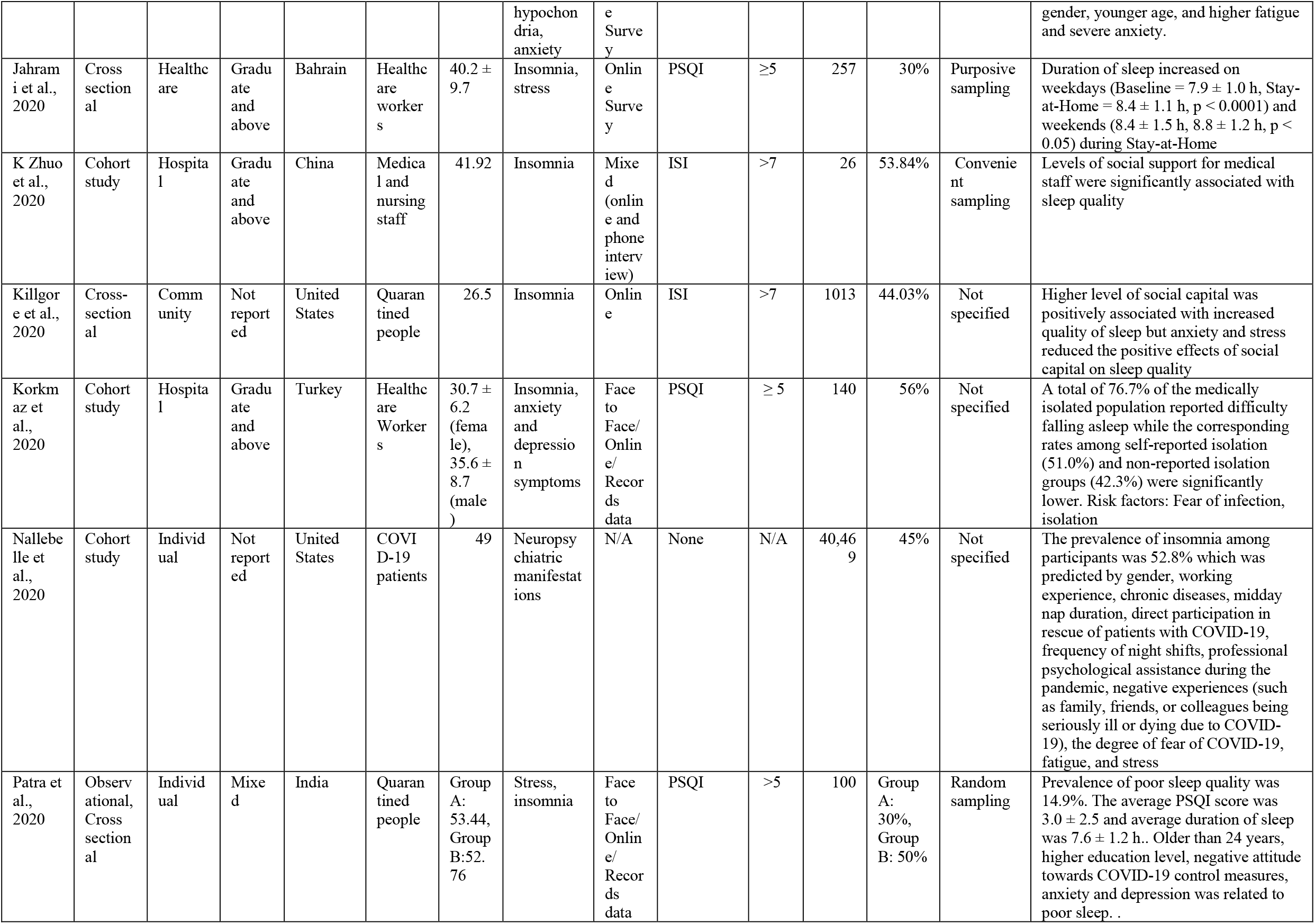
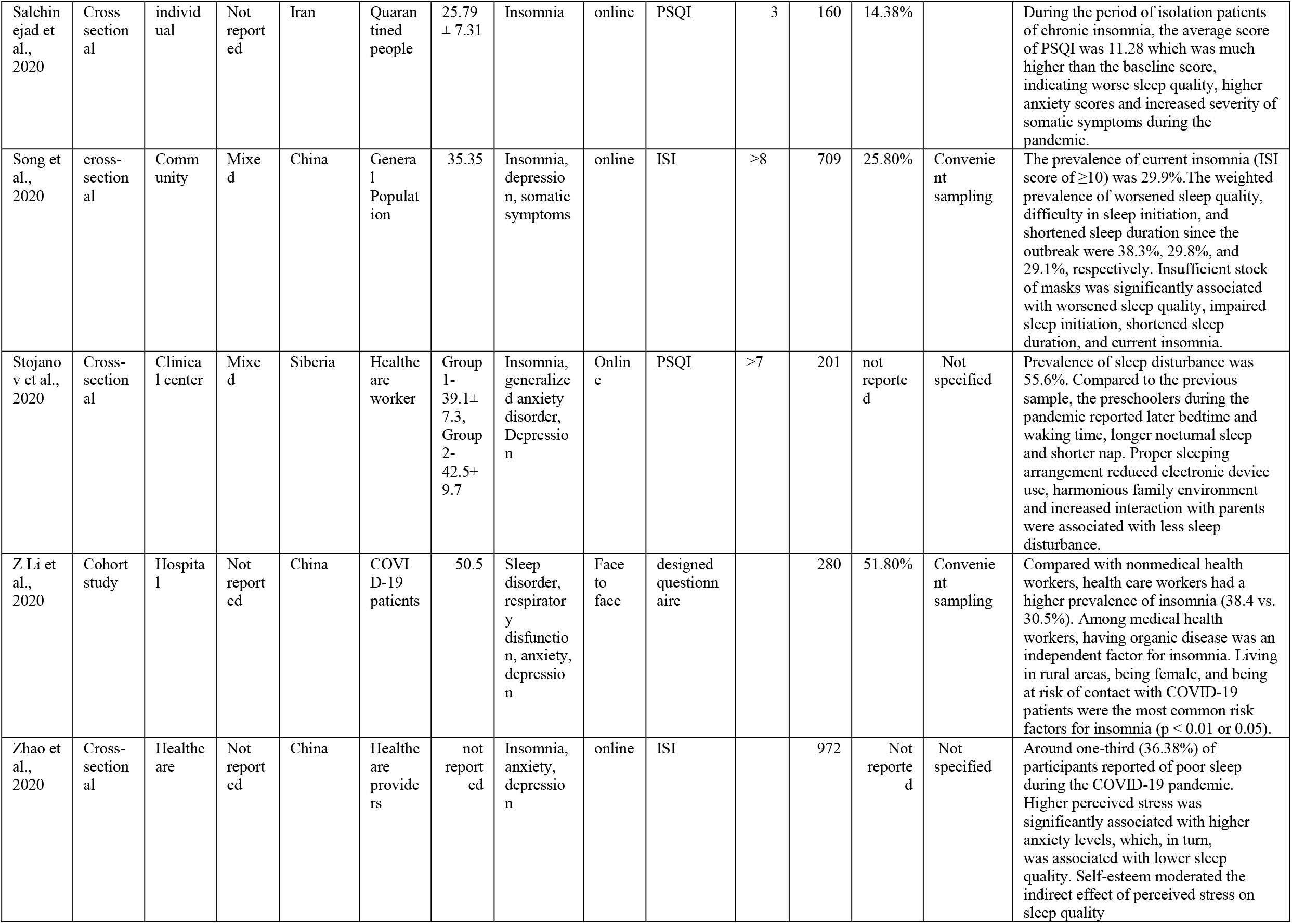
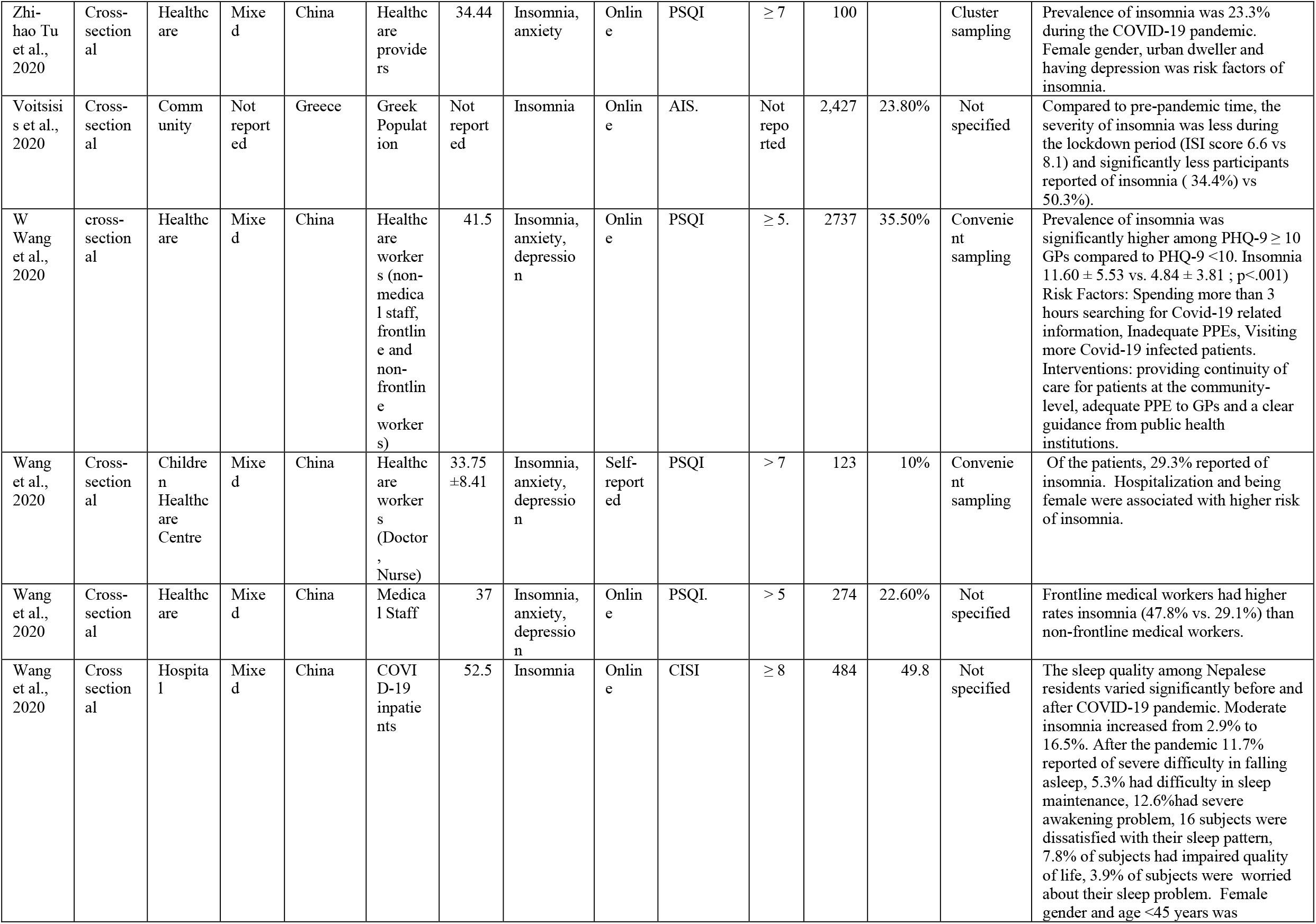
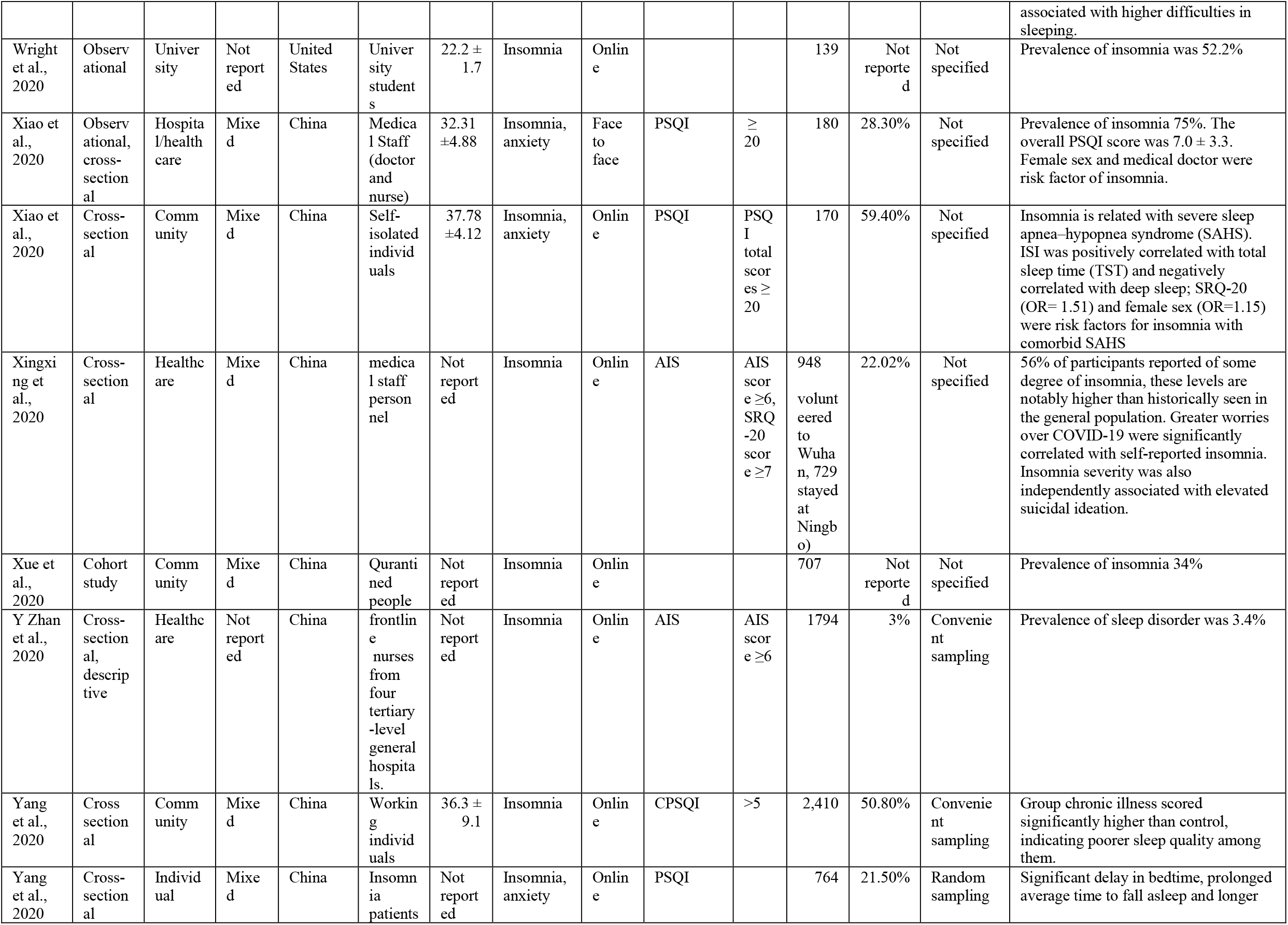
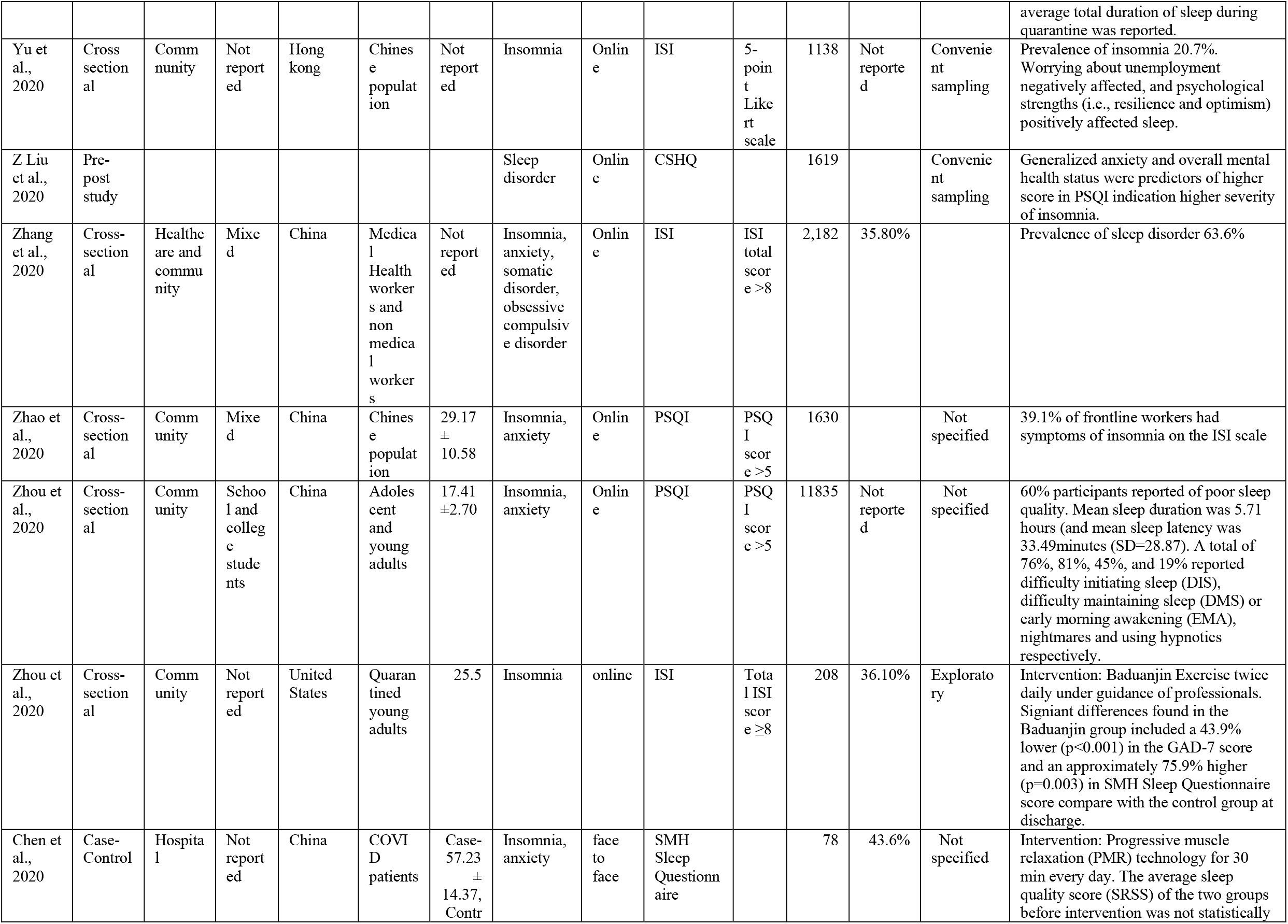
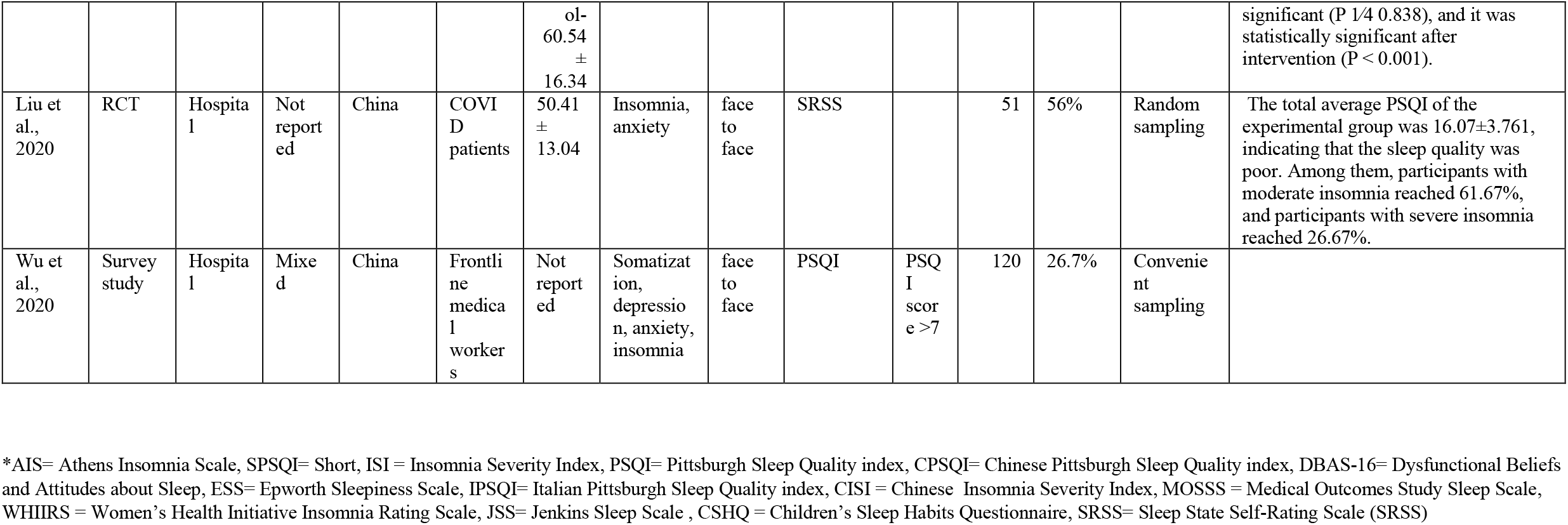

### 3.2 Characteristics of study population

#### 3.2.1 Education

Most studies did not report stratified levels of education among the participants or recruited people with mixed education status. In 16.25% (n = 13) of studies, the participants had at least bachelor’s degree. For example, according to Li et al.,^48^ 88.37% of the participants had bachelor’s degree. Only 20% (n = 16) of the studies included participants with graduate degree or above. According to Bhargava et al.,^49^ all the participants included in the study were specialized doctors who had graduate degrees or above. Furthermore, 26.25% (n = 21) studies recruited participants with various level of education. For example, Dai et al.,^50^ reported that, 60.69% of the recruited participants had undergraduate degree, 31.99% had graduate degree and 7.32% had high school degree.

#### 3.2.2 Population Specifics

Nearly one third (n = 22, 27.5%) studies were based on healthcare workers. For example, the study by Korkmaz et al.,^37^ was conducted among nurses, physician and other health care staff in the hospital. One fourth of the studies (n = 20) focused on general population. For example, Innocenti et al.,^46^ invited general populations of Italy to participate in their study. Alongside, 8.75% (n = 7) of the studies included only COVID-19 patients. Diomidous et al.,^39^ included participants (n = 204) from Greece who were confirmed cases of COVID-19. A few studies, (n = 6, 7.5%) included only quarantined people. For example, Salehinejad et al.,^51^ included participants who were quarantined at home during COVID-19.

#### 3.2.3 Age

Mean age ranged from 7.89 years to 63.9 years. Türkoğlu et al.,^45^ included 46 children with autism spectrum disorder and the mean age of the participants in this study was 7.89 years. Pinto et al.,^52^ a study based on Portugal population, included 365 participants in their study and the mean age of the participants was 63.9 years.

#### 3.2.4 Gender (% of males)

The total percentage of male participants in the study varied widely. It ranged from 3% to 82%, with a median value of 37.5%. Nevertheless, in majority of the studies the percentage of male participants were between 40%-60%. For example, Gupta et al.,^53^ reported of 58% male participants whereas, Li et al.,^54^ reported of 43% male participants.

### 3.3 Prevalence of sleep disorder

The prevalence of sleep disorders varied across the samples. Ranging from 2.3% to 76.6% For example, Zhang et al.,^55^ reported the overall prevalence of insomnia was as high as 76.6% among the medically isolated population. The prevalence rates varied across geographic regions of corresponding primary studies. For example, the prevalence of insomnia in the United States ranged between 20-41%. For example, 30% participants reported of some degree of insomnia in the study by Bhargava et al.,^49^ in the USA. Whereas, a significant variation was noted in the prevalence of insomnia in Chinese population. Although only 2.3% of the Chinese workforce reported of insomnia^56^, 58% of the frontline medical staff were suffering from insomnia^44^. Such variation was also noted in Italy, where the prevalence of insomnia ranged from 37.6% to 57%. For example, according to Casagrande et al.,^57^ 57.1% participants in Italy had insomnia and poor sleep quality.

### 3.4 Factors associated with sleep disorders during COVID-19

#### 3.4.1 Sex

20% of the studies (n =15) reported that females had a higher risk of some degree of insomnia. For example, Batool-Anwar et al.,^58^ reported that stratification by gender revealed worsening insomnia only among women. However, two studies also reported that males had higher risk of insomnia. For example, male participants in the study by Anzar et al.,^59^ reported higher prevalence of insomnia and poor sleep quality.

#### 3.4.2 Age

Only 7 studies reported that younger population had worse sleep compared to their older counterpart. For example. Beck et al.,^60^ mentioned that, younger population reported higher sleep disturbance compared to older population (79% vs 72%). Nevertheless, 4 studies reported of poor sleep among the older population as well. For example, Wang et al.,^61^ reported that older population had 1.42 times higher risk of insomnia.

#### 3.4.3 Education

Several studies found that level of education was associated with worse sleep. Zhang et al.,^62^ reported that education level of high school or below increased the risk of insomnia but ^33^ reported that participants having higher educational level had worse sleep.

#### 3.4.4 Social support

Social support was an important factor associated with sleep. Four studies reported that those with lack of social support had higher prevalence of insomnia and poor quality of sleep. For example, Tselebi et al.,^47^ reported participants with lack of family support had higher level of insomnia. Individuals in isolation^52^, living alone^63^ suffering from loneliness^64^ or being single ^44^ also reported of high degree of insomnia. This further emphasizes the impact of social support on sleep.

#### 3.4.5 Physical health

Individuals with preexisting chronic disease (n = 4) and perceived poor health (n = 5) suffered from higher prevalence of insomnia.

#### 3.4.6 Mental health

Insomnia worsened among patients with preexisting psychiatric illness (n = 4). Stress (n = 6), anxiety(n = 8), depression (n = 4), fatigue Zhang et al., ^62^ and low mood Carrigan et al.,^65^ was also found to be associated with poor sleep.

#### 3.4.7 Lack of Physical activity

Three studies reported that reduced opportunities of physical activities were associated with poor sleep among participants.

#### 3.4.8 Being Health Care Worker

Nearly 13% (n =10) of the studies reported that, health care workers were at risk of some degree of insomnia. For example, Anzar et al.,^59^ reported that HCW had not only a higher prevalence of insomnia but also had reduced quality of sleep.

Four studies reported that the health care workers at risk of contracting COVID-19 had poor sleep quality. For example, Carrigan et al.,^65^ found that, being at risk of CVID-19 at work lowers the quality of sleep. Zhang et al.,^62^also found that working in the isolation unit impaired the sleep cycle among HCWs. Frontline medical workers suffered from higher level of insomnia compared to non-frontline workers. For example, Qi et al.,^66^ reported that Frontline worker in the Wuhan, China (epicenter of the outbreak in China) scored significantly higher in the PSQI scale compared to non-front liners in Ningbo. Shift work ^28^, reluctance to join the frontline ^67^ and higher burden of work ^68^ also played an important role in sleeping.

#### 3.4.9 Factors Specific to COVID-19

Two studies reported that patients suffering from COVID-19 reported poor quality of sleep. Fear of COVID-19 (n = 2) and worry about the disease (n = 5) also affected the quality of sleep. Uncertainty due to the pandemic (n = 5), negative attitude toward control measures such as wearing mask ^69^ and lack of availability of testing ^50^ were the other factors specific to COVID-19 associated with sleep.

#### 3.4.10 Factors related to work

Several factors related to employment also impacted sleep significantly. One study found that people with low income had higher risk of sleep disorder compared to their high-income counterparts^33^. Worrying about impact of this pandemic on livelihood, possible negative impact on their income was also associated with sleep disorders^63^. One study mentioned that being at home without any work also impaired regular sleep cycle^52^. On the contrary, stress for returning to work after the lockdown did not impair sleep^56^.

### 3.5 Interventions for sleep disorders

There were only two interventions for sleep disorders. The study by Chen et al.,^70^ was conducted among COVID-2019 patients, where the intervention group performed Baduanjin exercise (traditional Chinese mind-body exercise routine) under professional guidance and the controls received usual care. Their anxiety and insomnia level was measured using GAD-7 and SMH Sleep questionnaire at baseline and discharge. Although there was no significant difference among these groups at baseline, intervention group showed 44% lower score (P<0.001) in GAD and 76% lower score (P=0.003) in the SMH Sleep questionnaire at discharge which indicated that Baduanjin exercise can be beneficial for improving anxiety and insomnia among COVID-19 patients.

In the randomized controlled clinical trial by Liu et al.,^36^ patients with COVID-19 patients who entered the isolation ward were randomly divided into experimental and control groups where the experimental group used progressive muscle relaxation (PMR) technology for 30 min per day for 5 consecutive days and the control group received only routine care and treatment. The Spielberger State-Trait Anxiety Scale (STAI) and Sleep State Self-Rating Scale (SRSS) were used to measure and record patient anxiety and sleep quality pre and post intervention. Both the group had comparable scores before intervention however, STAI and SRSS scores reduces significantly after intervention (P <0.001).

## 4. Discussion

### 4.1 Overview of the synthesized findings

After a thorough review of the available evidence, there were 78 studies that met the pre-set criteria of this scoping review. Most of the studies included were cross-sectional in design, and online questionnaires were the common mode of data collection. Studies were conducted both in the community and hospital settings. Most of the studies on sleep disorders were conducted in China or the USA. Insomnia, anxiety, and depression were commonly assessed disorders, and PSQI, ISIS, and AIS were used to evaluate insomnia and sleep quality. The findings from these existing literatures inform a high burden of sleep disorder during this pandemic. The prevalence of sleep disorder varied widely across the samples and ranged from 2.3% - 76.6%. The majority of the studies reported females had a higher risk of some degree of insomnia. Although older adults generally have a higher rate of insomnia, many studies have reported that younger generations were having difficulties with sleeping during the pandemic. Covariates-adjusted research was limited which could explain how age played a role in sleep patterns among the study samples. Moreover, the education level was also associated with sleep disorders, but there was ambiguity regarding the specific educational status. Both higher education and high school level education were found to impact sleep. This can be attributable to the fact the educated individuals or students may have academic and professional stressors that may have impacted their mental health and sleep conditions^71–73^. Furthermore, social support was found to have a critical role in sleep status and associated disorders. Lack of social support, family support, living alone, and isolation was associated with a higher risk of sleep disorders.

This review also found that comorbid physical and mental illness increased the risks of insomnia. Health care workers, especially those serving the frontline, were at high risk of insomnia during the COVID-19 pandemic. Increased workload, shift work, fear of contracting the virus were the significant risk factors among the HCW. This might have resulted in higher psychosocial stress and burnout, which can be associated with sleep disorders among the frontlines^74^. The fear of contracting the virus, worry about the disease, uncertainty regarding treatment and prevention measures, and negative attitude toward control measures were COVID-19 specific risk factors for sleep disorders. Other related factors were lack of opportunity for physical activity, fear of the negative impact on income, and staying at home without employment. Only two interventions were identified, which focused either on baduanjin exercise (as one of the traditional Chinese mind-body exercise) or progressive muscular reaction. Both reported significant improvement on sleep score after the intervention ^75,76^

The findings of this current review are comparable with the previous literature. Our review reports a considerably higher prevalence of both insomnia and insomnia symptoms higher during this pandemic. Several reviews have also reported higher prevalence after stressful events like stroke, chronic injuries^77^. The current review reports that females were at higher risk on insomnia and many studies have found that female are at high risk several mental illnesses such as anxiety and depression^78^.

Despite the lack of accurate normative data on evolution of sleep architecture with age, several studies suggest that sleep patterns of the individuals change across the lifespan. In older adults have a significant delay in sleep latency and decrease in total REM sleep^79^. These findings suggest that older adults have more problems with sleep compared to the younger population. However, several studies included in this review reported that younger population had more trouble with sleep. This could be explained by the preexisting and emerging academic and processional uncertainty and the mental stressors amid this pandemic.

Our findings are also in line with evidence suggesting that individuals with insomnia have a higher risk of psychiatric disorders and sleep disturbances may facilitate the onset of several mental health illness. Many studies have reported that there is a bidirectional relationship between anxiety, depression and insomnia^80^. In this circumstance high rate of insomnia among individuals with preexisting psychological was unsurprising.

Our review shed light on the high prevalence of insomnia and sleep disturbances among health care workers especially those in frontline. Studies from previous major outbreaks of SARS, MERS have also reported of elevation of psychological distress among HCW during and after the outbreak which interfered with their social and occupational functioning^81–83^. We need to learn from the history and intervene at the early stage to support these HCWs.

#### Limitations of the review

This scoping review has several limitations. The foremost limitation of the current literature is that most studies are cross-sectional or longitudinal design, most of which did not collect data before the onset of the pandemic to provide a comparative picture of the situation. The limited number of studies reporting prevalence at multiple time points made it difficult to determine how the insomnia status of individuals changed over time.

To investigate temporality and establish causal relationships, risk factors should be captured before the inception of the disease. Owing to this, causal relationship was not established. Therefore, the conclusion of this scoping review is limited to demonstrating association only.

Additionally, there is a risk of potential selection bias as we did not search all the databases and excluded unpublished studies, reports, and studies in languages other than English. Moreover, there could be a publication bias within the scientific literature as less significant findings are less likely to get published, thus may not contribute to the evidence base. Furthermore, we did not conduct a quantitative evaluation of the patient-level data, which could have eliminated within-study and between-study variations to provide uniform prevalence estimations across samples.

Another notable limitation of this review was limiting the search strategy within bibliometric databases, whereas a major proportion of the literature can be available on preprint servers. As we aimed to synthesize the peer-reviewed empirical research, we did not search the preprint sources that might have had early stage findings rather than peer-reviewed articles. Lastly, as most studies were conducted in China and the USA, our findings’ generalizability may be limited because of the wide variation of cultural norms and the healthcare system across the globe. Sleep disorders and co-morbid mental health problems may be highly prevalent in people living in low-income countries that are underrepresented in the existing studies^84,85^. Also, people with preexisting neuropsychiatric conditions or psychosocial vulnerabilities may experience a disproportionate burden of sleep disorders, that is yet to be examined through empirical research. Our review may not have captured the epidemiology of sleep disorders in those marginalized populations.

### 4.2 Implications for future research

The evidence landscape on COVID-19 and associated mental health problems has been changing rapidly^86,87^. This scoping review examined the epidemiological aspects of sleep disorders. As this review includes literature up to August 12, 2020, the synthesized evidence can be different from those studies published after that date, or those research that are being conducted now. However, as the goal of a scoping review is to chart the initial evidence in a scientific topic, the findings if this review can serve as the basis of future research focusing on specific subtopics of sleep disorders amid this pandemic. Such areas can be examined using specific methods appropriate for respective research objectives.

As most studies in this review were cross-sectional in nature, future studies should adopt longitudinal designs that may explore how the magnitude and correlators of sleep disorders evolve over time. Moreover, it is essential to examine the comorbid physical and mental health problems that are associated with sleep disorders (Hossain et al., 2020). Understanding the syndemic effects of multiple health problems may better inform why the prevalence and risk factors of sleep disorders may vary across samples^88,89^. Furthermore, it is essential to examine the mental health problems associated with sleep disorders in high risk populations such as those with neurodevelopmental disorders or social-economically marginalized populations. Evaluating the Epidemiology of sleep disorders in diverse population groups through cohort studies may provide valuable insights not only during this pandemic but also how different populations may experience unique psychosocial challenges during public health emergencies.

One of the notable findings of this review was the use of multiple scales for assessing sleep disorders as well as associated mental health problems. Many scholars have adopted previous scales, where is several new scales were introduced during this pandemic^90^. It is essential to conduct extensive factor analysis evaluating the appropriateness of multiple skills so that future research can be conducted using standardized scales and measurements. such efforts may provide a more accurate scenario of the epidemiology of sleep disorders.

The emergence of online data collection measures may provide easy access to people who may use those platforms; however, people with digital divide may not participate in such studies^91,92^. it is essential to consider such limitations of the existing research efforts and design inclusive methodology’s that may include diverse populations. In future research, measurements on psychosocial health outcomes including sleep disorders can be included in primary care and hospital health records that may offer valuable information on sleep disorders and other mental health problems. Nonetheless, Population based research exploring health and social aspects of COVID-19 may include instruments measuring the impacts of this pandemic on sleep behavior in different populations. Such approaches may improve the research production and the quality of data on Epidemiology of sleep disorders during this pandemic.

### 4.3 Implications for mental health policymaking and practice

This review examined the Epidemiology of sleep disorders, which may provide meaningful insights for mental health policy making and practice. The ongoing interventions and preventive measures adopted in different contexts should revisit the components that may influence sleep outcomes across populations. Institutional and local policies and programs on mental health should not only target mental disorders such as anxiety and depression, but also emphasize on sleep disorders that are highly prevalent in COVID-19 affected populations. Specific provisions should be made to prevent, diagnose, and treat sleep disorders. The emergence of telemedicine and online support systems provide unique opportunities from a public mental health perspective^93^. such services should be developed and implemented considering the burden and risk factors of sleep disorders during this pandemic. psychiatrists and psychologists may access over arcing risk and protective factors of sleep disorders and other mental health problems that can be managed using cost effective and evidence-based approaches. It is necessary to adopt such guidelines and services in psychosocial care policies and programs in different contexts. such measures can be customized using local mental health data considering the available mental health resources in those contexts. Furthermore, pandemic preparedness and mental health action plans should be informed by the epidemiological evidence on sleep disorders that may potentially impact the health and well being of health care providers, hospitalized patients, informal caregivers, and population at risk during COVID-19 pandemic.

Evidence-based pharmacological and psychosocial interventions have paramount importance for addressing sleep disorders. However, despite the elaborate searching through the databases, only two interventions were identified that focused on improving insomnia during the pandemic. Both of these interventions took place inside hospitals among patients admitted with COVID-19. Whereas, our review indicates a high prevalence of insomnia among patients, HCW as well as the general population. This necessitates community-based interventions to improve sleep among the target population. Mental health practitioners may access the relevance and appropriateness of existing interventions that may help during this pandemic. It is essential to conduct implementation research examining the effectiveness and efficiency of available interventions and develop newer interventions that may target specific psychosocial stressors for sleep disorders during this pandemic.

The findings of this review, emphasizes the need for early detection and effective treatment all the symptoms of insomnia, including the mild ones, before they evolve to more complex and evokes enduring psychological responses. Current knowledge of prevalence, type and comorbidities of prevalence, symptom profiles and comorbidities of insomnia should be utilized and tailored interventions targeting the behavioral components should be further developed to address the issue.

It has been evident that there is widespread of insomnia among the general population in this pandemic. All patients coming in contact with health care facilities should be screened, and those diagnosed with any form of insomnia should be referred to appropriate resources. As healthcare workers reported a higher burden of insomnia, specific interventions designed for addressing the problems at health care facilities should be available. The majority of health care workers are overburdened. If their optimal health and wellness are not ensured, their ability to work will be further compromised impose a severe predicament on the current fragile healthcare system.

Cognitive behavioral therapy for insomnia (CBT-I) is the treatment of choice for insomnia. A systematic review and meta-analysis of RCTs reported that, CBI-I is the effective mode of treatment for insomnia capable of producing clinically meaningful effect size with no identifiable adverse effects^94^. Several studies have also shown small to large effects on efficiency and quality and sleep onset latency along with reduction in severity of insomnia, wake after sleep onset and number of awakenings^95^. Moreover, a full economic evaluation of CBT-I in adult populations revealed CBT-I was cost-effective compared to pharmacotherapy or no treatment. Thus, incorporation of some form of CBT-I in clinical practice will improve the overall sleep status of the population.

If the delivery of in-person CBT-I is not possible due to the social distancing measures for preventing the spread of the virus, similar interventions can be delivered using the virtual platforms. CBT-I delivered though internet or computer was also found to be as effective and a viable alternative in the current context ^96–98^. Alongside, other avenues of telepsychiatry such as virtual clinics, remotely delivered psychotherapies, psychoeducation, 24/7 chat lines and digital monitoring should also be explored.

For areas with poor internet connection or individuals who are not accustomed with modern intervention formats, self-help books may be an effective alternative which could possibly reduce ensure standardization of care, quality control and optimal resource utilization^99^.

Accumulating evidence show that exercise and dietary interventions are associated with improved quality of sleep. A large RCT conducted in China reported that low fat diet and increased amount of exercise resulted in improved sleep by altering the metabolic pathways. Future intervention research should focus on how lifestyle-based interventions can impact sleep quality and overall mental health in different populations at risk. It is critical to assess the risk and protective factors that can be used to develop mental health promotion programs that potentially prevent sleep disorders among the vulnerable individuals. The scope of digital interventions, peer support groups community-based mental health services, self-management programs, And public mental health resources should be evaluated for promoting mental health and preventing sleep disorders across populations.

## 5. Conclusions

Amid this global pandemic burden of mental health issues are becoming a growing concern in addition to infection control. Sleep disorders are significant mental health problems associated with increased psychosocial stressors. Findings from this review suggest a high burden of sleep disorder across different population groups. Female gender, younger population, HCWs, COVID-19 related stressors were the major factors associated with sleep disorder identified in this review. Despite the high burden, a limited number of interventions were identified to address this problem. Early diagnosis of sleep disorder and adequate treatment is crucial to prevent further worsening of the condition. Evidence-based pharmacological and psychosocial interventions have paramount importance for addressing sleep disorders. Future studies should explore interventions that utilize digital platforms and adopt innovative strategies in order to ensure increased outreach and sustainability.

## Data Availability

All data related to this review are available from upon request.

## Acknowledgment

None

## Notes

Conflicts of interest: None

### Competing Interest Statement

The authors have declared no competing interest.

### Funding Statement

No funding was received to conduct this review.

### Author Declarations

This is a systematic scoping review that used publicly available published literature and did not involve any human participants. Therefore, this review did not require any IRB approval.

